# Probiotic effect of trichloroacetic acid on cervicovaginal microbiota in cervical intraepithelial neoplasia: A pilot study

**DOI:** 10.1101/2024.01.08.24301017

**Authors:** Takeo Shibata, Ayumu Ohno, Isao Murakami, Masahiro Takakura, Toshiyuki Sasagawa, Tadashi Imanishi, Mikio Mikami

## Abstract

Symbiosis of bacteria and human papillomavirus (HPV) in the cervicovaginal environment influences cervical intraepithelial neoplasia (CIN) progression or regression. In this case series, we enrolled all 10 patients who had resolved CIN after a cervical local treatment using trichloroacetic acid (TCA). Prominent changes in the cervicovaginal microbiota, such as an enrichment of the genus *Bifidobacterium* and genus *Lactobacillus*, were observed in seven of 10 patients. A decrease in cervicovaginal bacterial alpha diversity was observed in 4 patients with high-risk HPV clearance. Differential abundance analysis revealed that genus *Bifidobacterium* increased significantly after TCA. The dominance of a single bacteria can be characteristic of CIN cures after TCA. These results highlight the potential link of cervicovaginal bacteria such as genus *Bifidobacterium* and genus *Lactobacillus* in the clearance of CIN and high-risk HPV. This pilot study guides future research questions that specific cervicovaginal bacteria may be promising candidates for probiotic therapy to treat CIN and HPV infections.

## Introduction

About 5% of all cancers are attributed to human papillomavirus (HPV) infection ***Forman et al.*** (***2012***). HPV infects mucous membranes and causes a variety of diseases such as genital warts, precancerous cervical lesions (cervical intraepithelial neoplasia: CIN), cervical cancer, vaginal intra-epithelial neoplasia (VAIN), vaginal cancer, vulvar cancer, penile cancer, anal cancer, head and neck cancer, and even recurrent respiratory papillomatosis throughout the body ***Doorbar et al.*** (***2015***) ***WHO*** (***2021***). HPVs are classified into more than 200 genotypes, some HPVs are categorized based on the risk of developing cancer, such as high-risk HPV (HPV types 16, 18, 31, 33, 35, 39, 45, 51, 52, 56, 58, 59, 66, and 68) ***Doorbar et al.*** (***2012***) ***Hum*** (***2021b***). Especially, HPV type 16 or HPV type 18 causes 70% of cervical cancers and CIN ***Hum*** (***2021a***). The development of HPV-related diseases, namely CIN and cervical cancer, has been elucidated in considerable detail in molecular biology ***Doorbar et al.*** (***2012***). E6 and E7 proteins as viral oncogenes and proteins are key to carcinogenesis. They modulate the activity of p53 and retinoblastoma family proteins ***Doorbar et al.*** (***2012***).

The factors that influence the progression or regression of CIN are unknown, however, HPV is the evident cause of CIN ***Sasagawa et al.*** (***1992***) and cervical cancer ***Pal and Kundu*** (***2020***). HPV infection is a common infection that affects more than half of the cases within 5 years of the onset of sexual intercourse ***Winer et al.*** (***2003***). However, the incidence of cancer is low compared to the infection rate of HPV ***Winer et al.*** (***2003***). Evidence suggests an association between symbiotic bacteria in humans and carcinogenesis ***Doocey et al.*** (***2022***). The cervicovaginal bacterial community have been shown to be associated with the progression or elimination of CIN or HPV infection ***Mitra et al.*** (***2016***) ***Kyrgiou et al.*** (***2017***) ***Norenhag et al.*** (***2020***) ***Lin et al.*** (***2020***) ***Cheng et al.*** (***2020***) ***Usyk et al***. (***2020***). As a feature of cervical or vaginal microbiome (cervicovaginal microbiome), *Lactobacillus* are dominant, however, non–*Lactobacillus*-dominance or *Lactobacillus iners* are dominant in the patients with CIN or high-risk HPV positive ***Norenhag et al.*** (***2020***)***Lin et al.*** (***2020***). Non-*Lactobacillus*-dominant environment such as bacterial vaginosis (BV) was associated with higher rates of HPV infection, high-risk HPV persistence, and delayed HPV clearance ***Kyrgiou et al.*** (***2017***). In particular, cervicovaginal bacterial composition under HPV infection was characterized by BV-associated bacteria, which is an anaerobic bacteria such as *Sneathia, Prevotella*, and *Megasphaera* ***Cheng et al.*** (***2020***). In addition, *Gardnerella*-enrichment is associated with developing high-grade squamous intraepithelial lesion (HSIL) in a longitudinal study ***Usyk et al.*** (***2020***). Multiple HPV infections compared to no HPV infections were attributed to the increased bacterial biodiversity (i.e., high alpha diversity)***Cheng et al.*** (***2020***).

A classic excision procedure called cervical conization for CIN, which removes the cervix, has been reported to increase Obstetric adverse events such as preterm birth and premature rupture of membranes ***Bevis and Biggio*** (***2011***). To develop minimally invasive treatment, we conducted clinical trials for trichloroacetic acid (TCA) or phenol treatment on the CIN or VAIN lesions ***Maehama et al.*** (***2023***). In phenol-treated patients, disease regression rates after 24 months for CIN1-3 were 99%, 98%, and 100%, respectively ***Maehama et al.*** (***2023***). We observed that most patients resolved CIN or VAIN lesions upon TCA application within one year (in preparation). This chemical peeling method presumes to remove the epithelial lesions of such HPV-related diseases. The host immune responses are likely to be involved with the complete clearance of diseases by this method. TCA treatment can potentialy avoid cervical conization and hysterectomy and is a more desirable treatment for women who wish to bear children in the future.

We hypothesize that cervicovaginal bacterial dynamics are involved with the clearance of CIN and HPV infection after TCA treatment. In this pilot study, we investigated the difference in cervicovaginal bacteria before and after chemical peeling treatment by TCA in patients who had cleared not only CIN/VAIN but also high-risk HPV with TCA treatment. We attempt to identify bacteria that are candidates for probiotic treatment to establish safe and effective invasive treatment for CIN or VAIN.

## Results

### Patient characteristics and clearance of HPV

The average age of patients enrolled in this pilot study was 35.7 ± 8.1 years old (10 patients). The average TCA treatment duration of patients with CIN or VAIN lesions from initial sample collection to eventual cure was 289 days. The 10 patients with CIN (nine patients: one CIN1; two CIN2; six CIN3) or VAIN (one patient, VAIN2) diagnosed by colposcopy and biopsy were enrolled in this analysis (Table 1). The distribution of race or ethnicity was 100% Japanese. In pre-TCA treatment phase, cervical cytology was; atypical glandular cells: AGC (n = 1), adenocarcinoma in situ: AIS (n = 1), atypical squamous cells of undetermined significance: ASCUS (n = 1), high grade squamous intraepithelial lesion: HSIL (n = 4), and low grade squamous intraepithelial lesion: LSIL (n = 3). High-risk HPV was 90% positive before TCA treatment and 60% after TCA treatment (Table 1). However, at follow-up after TCA treatment we observed that high-risk HPV disappeared in all patients within 9 months (Table 2).

**Table 1.** Patient Characteristics.

| Test | Diagnosis | Number of Patients | Frequency (%) |
| --- | --- | --- | --- |
| Before Treatment |  |  |  |
| Cytology | AGC | 1 | 10 |
|  | AIS | 1 | 10 |
|  | ASCUS | 1 | 10 |
|  | HSIL | 4 | 40 |
|  | LSIL | 3 | 30 |
| Biopsy | CIN1 | 1 | 10 |
|  | CIN2 | 2 | 20 |
|  | CIN3 | 6 | 60 |
|  | VAIN2 | 1 | 10 |
| High-risk HPV* | Negative | 1 | 10 |
|  | Positive | 9 | 90 |
| After Treatment |  |  |  |
| Cytology | NILM | 10 | 100 |
| High-risk HPV* | Negative | 4 | 40 |
|  | Positive | 6 | 60 |

\*: Total 14 high-risk HPV genotypes were defined as HPV16, 18, 31, 33, 35, 39, 45, 51, 52, 56, 58, 59, 66, and 68. AGC: atypical glandular cells; AIS: adenocarcinoma in situ; ASUCUS: atypical squamous cells of undetermined significance; HSIL: high grade squamous intraepithelial lesion; LSIL: low grade squamous intraepithelial lesion; CIN: cervical intraepithelial neoplasia; VAIN: vaginal intraepithelial neoplasia; HPV: human papillomavirus; NILM: negative for intraepithelial lesion or malignancy.

**Table 2.** Clearance of HPV after TCA treatment.

| Cases | Diagnosis | HPV Before TCA | HPV After TCA | HPV at Follow-up After TCA |
| --- | --- | --- | --- | --- |
| Case 01 | CIN3 | 52 | 67 | Not examined* |
| Case 02 | CIN3 | 16 | 66 | Negative |
| Case 03 | CIN2 | 6, 16, 53, 54, 58, 66 | Negative | Negative |
| Case 04 | CIN2 | 31 | 31, 51 | Negative |
| Case 05 | CIN1 | 51, 82 | Negative | Not examined* |
| Case 06 | VAIN2 | 6, 62, 81 | 59 | Negative |
| Case 07 | CIN3 | 16, 51, 56, 59, 66, 70 | 59 | Negative |
| Case 08 | CIN3 | 16 | 52, 67 | Negative |
| Case 09 | CIN3 | 16, 52 | 58 | Negative |
| Case 10 | CIN3 | 16, 31 | Negative | Not examined* |

\*: No HPV typing was done at the time of follow-up for patients who were negative for high-risk HPV after TCA treatment. HPV: human papillomavirus; TCA: trichloroacetic acid; CIN: cervical intraepithelial neoplasia; VAIN: vaginal intraepithelial neoplasia.

### Statistics of reads and taxa

After importing 20 raw fastq files, we obtained a total of 1,812,889 reads, mean 90644.4 reads, median 83714.5 reads, minimum 6658 reads, and maximum 191,622 reads. After final quality filtering using q2-dada2 and q2-quality-control, we obtained 494 features (taxa) and a total of 714,869 features (taxa) in 20 samples. For read-length, the minimum length was 38, maximum length 283, and mean length 236.96.

### Characteristics of cervicovaginal microbiota pre- or post-TCA treatment

Among the identified bacteria, the top genera before and after TCA treatment are shown in Figure 1A. These top bacteria are mentioned below based on their frequency (%).

1. Genus *Lactobacillus*, after TCA, 38.9%
2. Genus *Lactobacillus*, before TCA, 37.3%
3. Genus *Gardnerella*, before TCA, 21.9%
4. Genus *Bifidobacterium*, after TCA, 16.4%
5. Genus *Prevotella*, before TCA, 12.9%
6. Genus *Gardnerella*, after TCA, 11.4%
7. Genus *Prevotella*, after TCA, 11.1%
8. Genus *Megasphaera*, before TCA, 7.14%
9. Genus *Atopobium*, before TCA, 6.87%
10. Genus *Atopobium*, after TCA, 4.19%
11. Genus *Megasphaera*, after TCA, 3.64%

**Figure 1.**
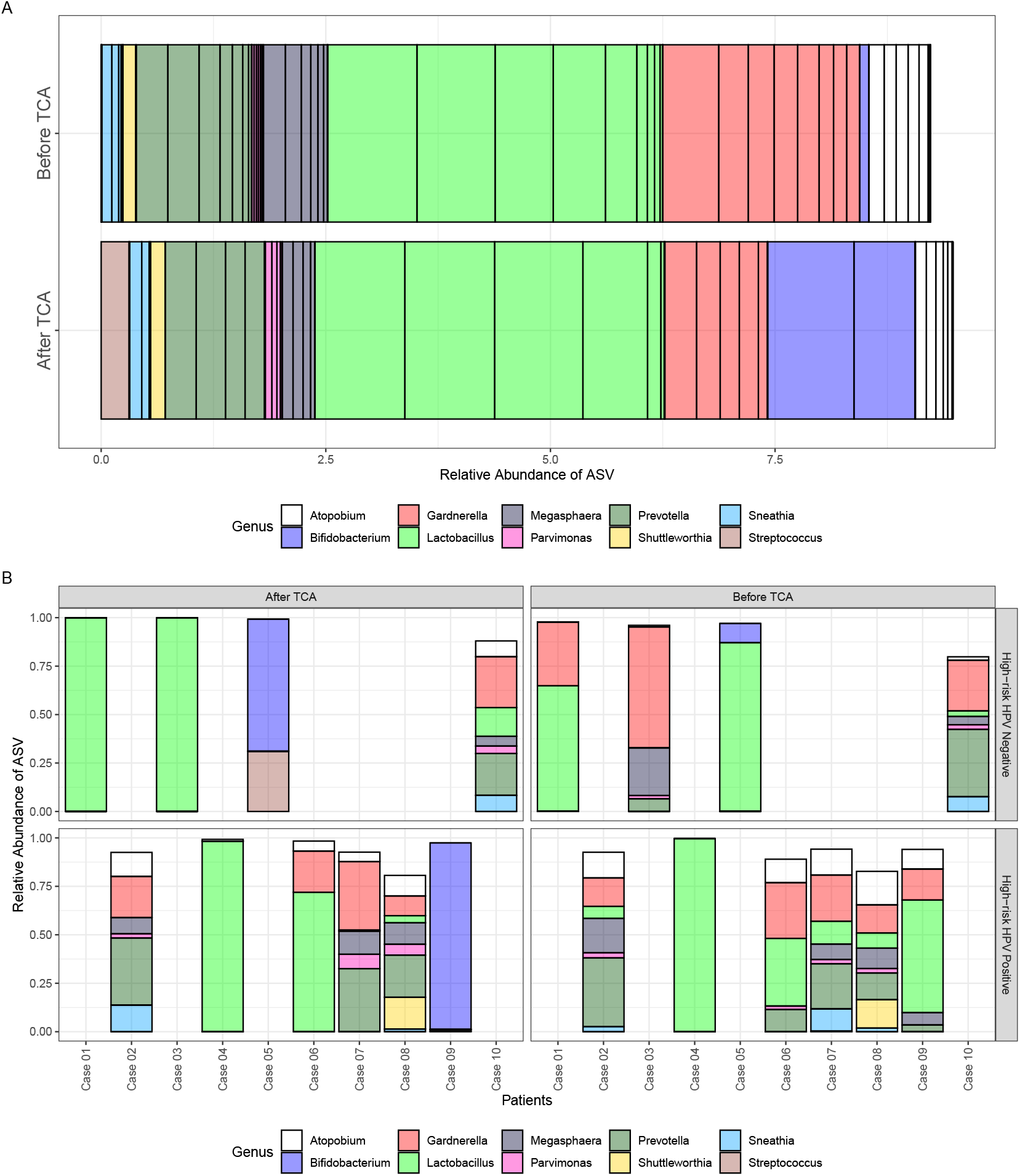
Characteristics of cervicovaginal microbiota before and after TCA treatment for CIN/VAIN. A: The relative abundance of taxa before and after TCA treatment is shown on the x-axis and the top 10 bacteria at the genus level are plotted in colored bar plots. B: The relative abundance of taxa is displayed for each patient, before and after TCA treatment, and with high-risk HPV clearance information after TCA treatment. TCA: trichloroacetic acid; ASV: amplicon sequence variant; HPV: human papillomavirus.

The frequency of the top bacteria, except for genus *Bifidobacterium* did not largely change before and after TCA treatment, for example, genera *Lactobacillus* (37.3% before vs. 38.9% after TCA treatment), *Gardnerella* (21.9% before vs. 11.4% after TCA treatment), *Prevotella* (12.9% before vs. 11.1% after TCA treatment), *Atopobium* (6.87% before vs. 4.19% after TCA treatment), and *Megasphaera* (7.14% before vs. 3.64% after TCA treatment). Contrarily, the genus *Bifidobacterium* changed significantly before and after TCA treatment; the frequency increased from 1% before TCA treatment to 16.4% after TCA treatment. Figure 1B shows the change in the cervicovaginal bacterial microbiota before and after TCA treatment for each patient. Figure 1B provides information on high-risk HPV clearance and persistence after TCA treatment. An enrichment of genus *Bifidobacterium* and genus *Lactobacillus* were observed in seven of 10 patients (Cases 01, 03, 04, 05, 06, 09, and 10) after TCA treatment. We found that genus *Bifidobacterium* (blue bar) increased in two patients (Cases 05 and 09) after TCA treatment. As an additional finding, in four patients (Cases 01, 03, 04, and 06), cervicovaginal bacterial microbiota was dominated primarily by the genus *Lactobacillus* (green bar, 99.9%, 100%, 98.2%, and 72.0%, respectively) after TCA treatment.

### Alpha and beta diversity analyses

Decline in alpha diversity was observed in seven of 10 patients (Cases 01, 03, 05, 06, 07, 09, and 10. Figure 2). Alpha diversity was consistently decreased in four patients with high-risk HPV clearance after TCA treatment (Cases 01, 03, 05, and 10, top panel of Figure 2). Both decreased alpha diversity (three patients, Cases 06, 07, and 09) and increased (three patients, Cases 02, 04, and 08) were observed in patients with high-risk HPV persistence after TCA treatment (bottom panel of Figure 2).

**Figure 2.**
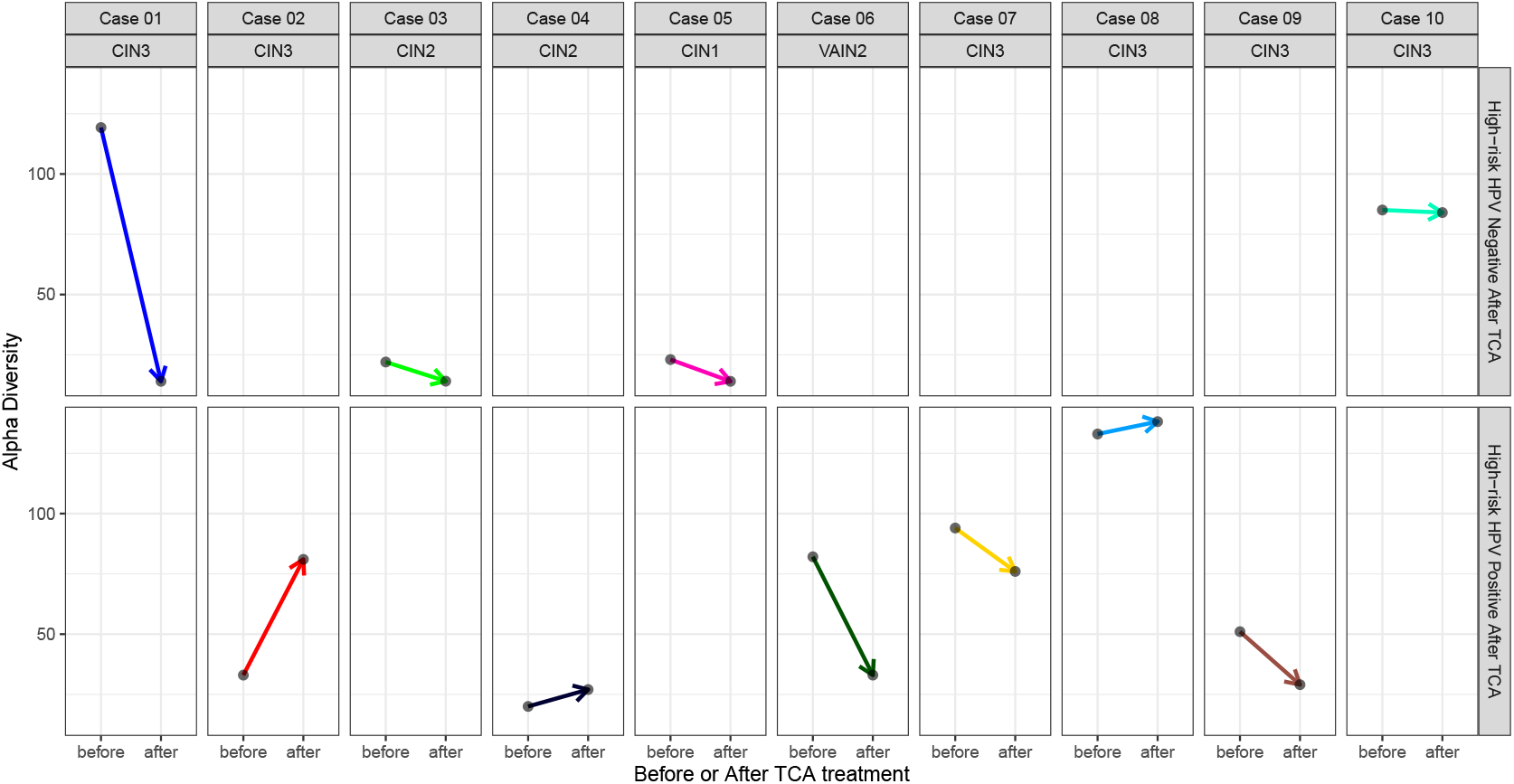
Analysis of alpha diversity before and after TCA treatment. Alpha diversity is reduced in high-risk HPV-negative cases, whereas alpha diversity is variable in high-risk HPV-positive cases. Alpha diversity was consistently decreased in four patients with high-risk HPV clearance after TCA treatment (Cases 01, 03, 05, and 10, top panel). In patients with high-risk HPV persistence after TCA treatment (bottom panel), both decreased alpha diversity (Cases 06, 07, and 09) and increased (Cases 02, 04, and 08) were observed.

We examined whether the cervicovaginal bacterial microbiota is affected by factors including; factor 1: individuals, factor 2: pre- or post-TCA treatment, and factor 3: high-risk HPV. Individual differences, not TCA treatment or high-risk HPV, define the similarity of bacterial composition in the cervicovaginal bacterial microbiota (Figure 3). About 50.9% of microbial variation could be explained by patient differences (R2 = 0.509, p = 0.030, Adonis test, Figure 3A). High-risk HPV infection before TCA treatment explained 6.8% variation of microbe composition (R2 = 0.068, p = 0.149, Adonis test, Figure 3C). High-risk HPV infection after TCA treatment explained 6.7% variation of microbe composition (R2 = 0.067, p = 0.131, Adonis test, Figure 3D). Before and after chemical peeling using TCA explained only 3.6% variation of microbe composition (R2 = 0.036, p = 0.408, Adonis test, Figure 3B).

**Figure 3.**
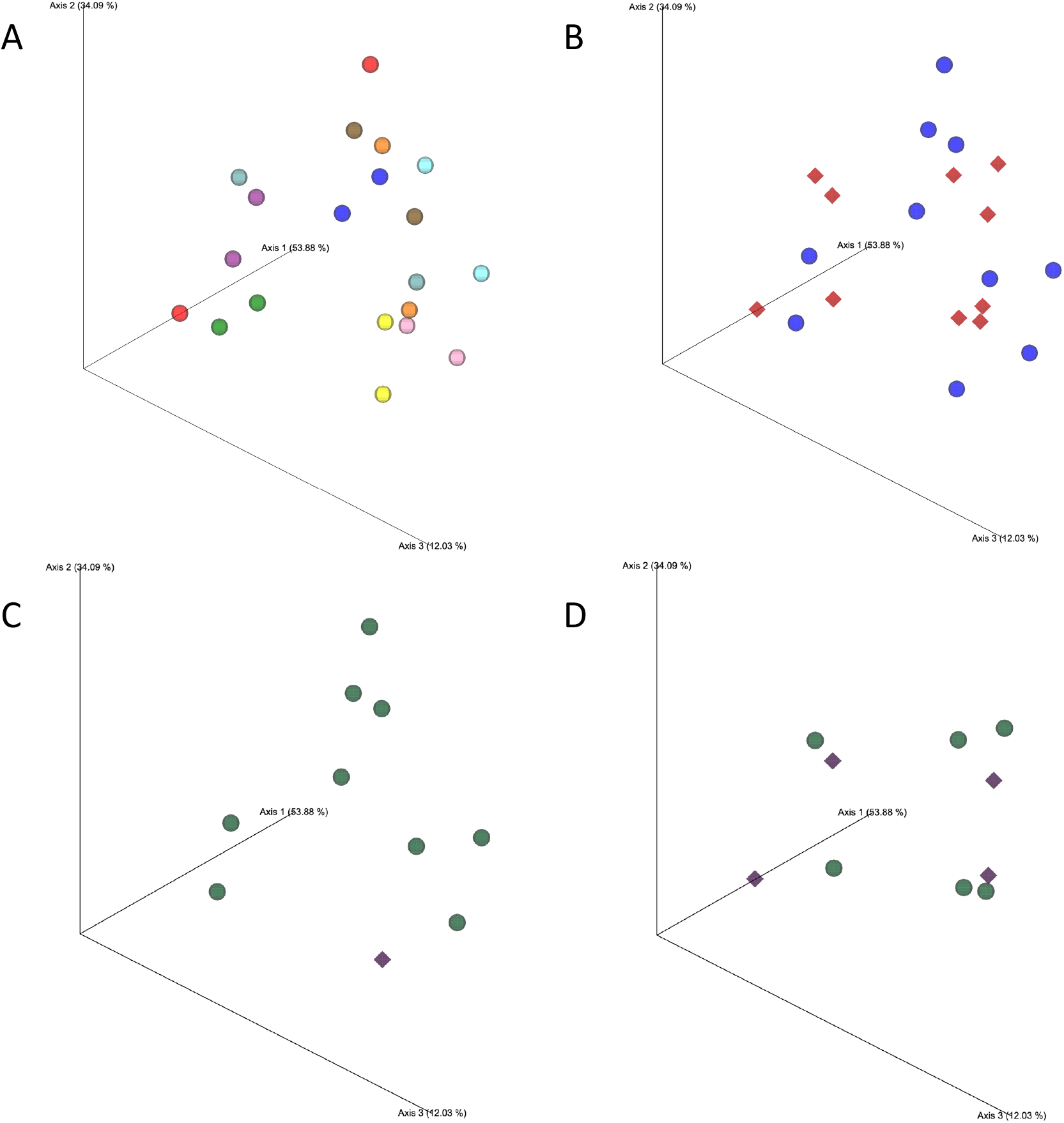
Principal component analysis (beta diversity) by patients, TCA treatment, and high-risk HPV. Samples are predominately clustered by (A) individuals. Samples are not clustered by (B) pre- or post-TCA treatment, (C) high-risk HPV infection before TCA treatment, and (D) high-risk HPV infection after TCA treatment. A: Samples are colored with 10 kinds of color for each patient. B: Samples are colored by after TCA treatment (red, diamond) and before TCA treatment (blue, circle). C: Samples before TCA treatment are colored by high-risk HPV positive (green, circle) and negative (purple, diamond). D: Samples after TCA treatment are colored by high-risk HPV positive (green, circle) and negative (purple, diamond). About 50.9% of microbial variation could be explained by patient differences (R2 = 0.509, p = 0.030, Adonis test, A). Before and after chemical peeling using TCA explained only 3.6% variation of microbe composition (R2 = 0.036, p = 0.408, Adonis test, B). High-risk HPV infection before TCA treatment explained 6.8% variation of microbe composition (R2 = 0.068, p = 0.149, Adonis test, C). High-risk HPV infection after TCA treatment explained 6.7% variation of microbe composition (R2 = 0.067, p = 0.131, Adonis test, D).

### Differential abundance analysis for cervicovaginal microbiota between pre- and post-TCA

There was a remarkable increase in genus *Bifidobacterium* in Cases 05 and 09 after TCA treatment, as shown in the bar graphs (Figure 1B). Consistent with this result at the genus level, we observed an increase in one *Bifidobacterium sp*. after TCA treatment, although we were unable to identify the species due to the resolution limitation of 16S rRNA analysis (log fold-change: −25.0, adjusted p-value < 0.001, Figure 4). Results for the genus *Lactobacillus* and *Lactobacillus sp*. were inconsistent between pre- and post-TCA treatment. Thus, some *Lactobacillus* species were enriched after TCA treatment, while others were before TCA treatment. The *Lactobacillus sp*. enriched before TCA treatment were not identified due to the resolution limitation of 16S rRNA analysis (log fold-change: 23.1, adjusted p-value < 0.001, Figure 4).

**Figure 4.**
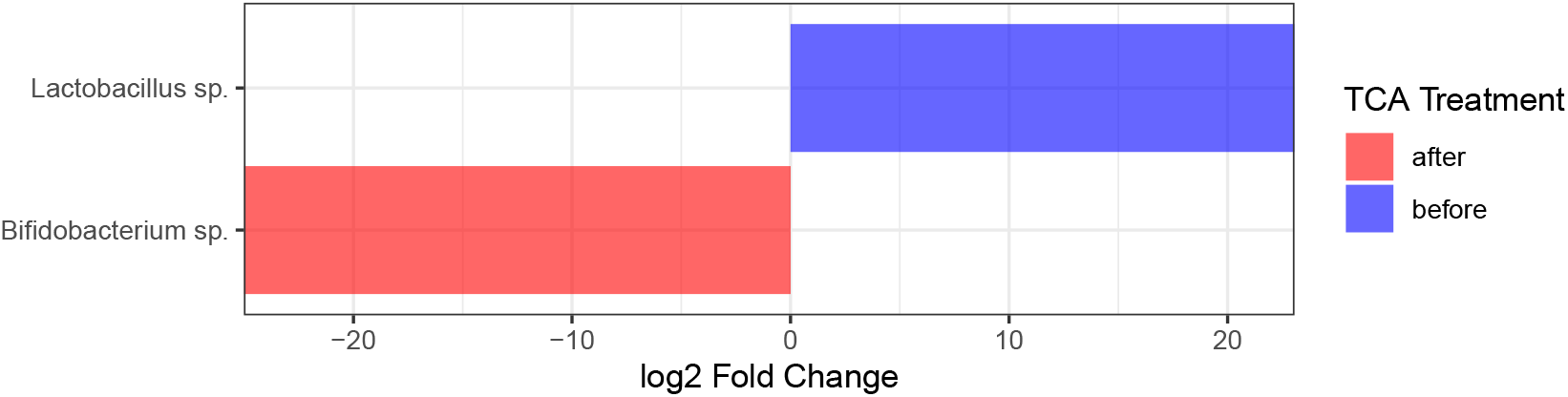
Enriched taxa pre- and post-TCA. *Bifidobacterium sp*. was detected in enriched taxa after TCA treatment (log fold-change: -25.01414, p-value: 2.687858e-17, adjusted p-value: 9.649411e-15). *Lactobacillus sp*. was detected in enriched taxa before TCA treatment (log fold-change: 23.06017, p-value: 6.417715e-15, adjusted p-value: 1.151980e-12)

## Discussion

Nonsurgical treatment of CIN should be especially considered for women of childbearing age. The effectiveness of TCA treatment for CIN has been analyzed not only by us but also by multiple research groups, and it may become a standard treatment for CIN in the future***Schwameis et al.*** (***2022***). However, there is a lack of data regarding the healing mechanism of CIN after TCA treatment. As represented in *Döderlein’s bacillus* described by Dr. Albert Döderlein, a relatively simple composition of bacteria such as *Lactobacillus* in the vagina is healthy. Persistent HPV infection increases the risk of developing cervical cancer and CIN; however, it is unclear what type of cervicovaginal bacteria are involved in the disappearance of CIN. This research is an attempt to analyze the relationship between cervicovaginal microbiota, high-risk-HPV, and the regression of CIN.

In this study, the mean duration required for the clearance of CIN or VAIN lesions after TCA administration was 289 days. After nearly a year of long-term follow-up along with high risk-HPV disappearance, it was observed that the cervicovaginal microbiota in CIN/VAIN-cured patients changed into an environment dominated largely by a single bacteria such as genus *Lactobacillus* or *Bifidobacterium* (Figure 1). Then, BV-associated bacteria such genera *Gardnerella, Prevotella, Atopobium*, and *Megasphaera* were decreased after TCA treatment (Figure 1). A simplification of the bacterial composition, suggested by a decrease in alpha diversity was observed in all patients with all CIN clearance and high-risk HPV negative, whereas an increase in alpha diversity was seen in patients with persistent HPV, suggesting that the cervicovaginal microbiome may influence the action of high-risk HPV elimination (Figure 2). The enrichment of *Bifidobacterium sp*. after TCA treatment in the patients with CIN or VAIN clearance is enough to be detected by differential abundance analysis, suggesting that the transformation into a single microbiota in the cervix and vagina, not only by *Lactobacillus* but also by *Bifidobacterium* may be the key to clearance of CIN or VAIN lesions (Figure 4). Taken together, this study infers the natural history of CIN clearance processes with TCA treatment associated with high-risk-HPV and the cervicovaginal microbiome. Thus, below is a possible mechanism by which TCA treats CIN; TCA first causes detachment of the CIN/VAIN lesion (phase I), changes in the cervicovaginal microbiome (phase II, increase in genera *Lactobacillus* and *Bifidobacterium* and decrease in BV-associated bacteria), clearance of high-risk-HPV (phase III).

We should be cautious about whether *Bifidobacterium* could be a candidate for probiotic treatment of CIN and HPV infections. However, previous studies suggest that *Bifidobacterium* is a useful bacterium against HPV infection. Mei et al. reported that *Bifidobacterium* were enriched in the patients with clearance of HPV 16 compared to patients with no history of high risk-HPV infection ***Mei et al.*** (***2022***). Similarly, this phenomenon was observed in Case 09 (i.e., HPV16 was negative at folllow-up after TCA treatment). Cha et al. reported the probiotic effect of *Bifidobacterium* on SiHa cervical cancer cell line expressing HPV 16 and found the suppression of E6 and E7 ***Cha et al.*** (***2012***). Further investigation is needed to decipher the relationship between *Bifidobacterium* and CIN and HPV persistent/clearance.

When discussing whether *Lactobacilli* affects the clearance of CIN and HPV, we should discuss it at the species level rather than the genus level. Ou et al. reported the effects of oral administration of *Lactobacillus rhamnosus* and *Lactobacillus reuteri* on HPV clearance and cervical cytology: rate of ASCUS and LSIL dropped from 21.0% to 6.5% in probiotic group vs 15.3% to 10.2% in control group, p = 0.017 ***Ou et al.*** (***2019***). According to Nicolò et al., *Lactobacillus gasseri* or *Lactobacillus jensenii* induce interferon-γ and these species may have the potential to assist immune cells with HPV clearance ***Nicolò et al.*** (***2021***). On the contrary, Brotman et al. showed that *Lactobacillus iners* had relatively higher proportion of HPV-positive samples, suggesting their link with the risk of HPV persistence ***Brotman et al.*** (***2014***). In this study, an increase in genus *Lactobacillus* was observed after CIN cure (Figure 1), but upon differential abundance analysis, *Lactobacillus sp*. decreased (Figure 4). That is, some *Lactobacillus spp*. increased before TCA treatment, while other *Lactobacillus spp*. decreased after TCA treatment. Analysis using long reads is necessary to identify the abundance at species level.

However, there are certain limitations of this study. In this case series, we analyzed only 10 patients with cured CIN, with emphasis on the homogeneity of the patient background. It is difficult to draw general conclusions from case series, therefore, in the next project that expands on this research, we plan to conduct a case-control study and time-series analysis of a large number of healthy women as well as CIN patients receiving TCA treatment. Their use in probiotic therapies for CIN require detailed basic microbiological studies and prospective clinical trials that were beyond the scope of this study. A clear understanding of the interplay between several factors, including oncogenic HPV, humans, bacteria, and possibly a fungus in order to control probiotic treatment that manipulates the composition of cervicovaginal bacteria is necessary.

The cervicovaginal microbiome is an important factor in the clearance of CIN and HPV. Low invasive local treatment with chemical peels using TCA may be associated with changes in *Lactobacillus* or *Bifidobacterium* in the cervicovaginal microbiota. This pilot study leads to the hypothesis that certain bacteria that fluctuate in the cervicovaginal ecology are promising as they may be prospective candidates for probiotic treatment to treat CIN and HPV infection. We advocate the importance of CIN management in order to understand not only the state of oncogenic HPV infection but also the cervicovaginal microbiome. Further intensive research in metabolome or immunity research is needed to examine both the role of the cervicovaginal microbiota in cancer and the use of probiotics for cancer treatment.

## Methods and Materials

### Ethics approval

This study was approved by the Institutional Review Board at the Kanazawa Medical University (IRB #I322). Written informed consent was obtained from all study patients.

### Patient selection and chemical peeling treatment with TCA for CIN and VAIN

Eligible patients who met the following conditions were selected for this pilot study; 1: Patients treated by TCA at Kanazawa Medical University during the period from 2017 to 2020; 2: A cytology specimen prior to TCA treatment was archived; 3: All patients were those who were cleared with CIN or VAIN after TCA treatment. Under the above selection conditions, 14 patients were treated with TCA, 12 patients had samples available, and 10 patients with cured CIN or VAIN lesions were included in the analysis. All 10 patients were diagnosed with CIN or VAIN using the punch biopsy specimens under the guidance of colposcopy. No patients have undergone a hysterectomy or loop electrosurgical excision procedure (LEEP) before or after TCA treatment. TCA (70%) was applied to wide areas of the cervical or vaginal lesions three to five times using a small cotton tip every 4 weeks. The Pap test using the liquid-based cytology (LBC) sample was performed in all patients every visit at 4 weeks intervals until disease clearance. Confirmation of CIN and VAIN lesion disappearance in colposcopy after Pap test-negative (i.e., negative for intraepithelial lesion or malignancy: NILM) was the definition of disease clearance. We examined cervicovaginal microbiota in the LBC samples before (CIN/VAIN lesion persistence) and after (CIN/VAIN lesion clearance) TCA treatment in this study.

### HPV genotyping

HPV-DNA was detected by Uniplex E6/E7 PCR method or GENOSEARCH™ HPV 31 (Medical & Biological Laboratories, Co., Ltd., Tokyo) which can detect up to 39 or 31 HPV genotypes each using LBC samples (ThinPrep solution, HOLOGIC, Marlborough, MA USA) ***Okodo et al.*** (***2018***). A total of 14 types including HPV16, 18, 31, 33, 35, 39, 45, 51, 52, 56, 58, 59, 66, and 68 were classified as high-risk HPV genotypes ***Campos-Romero et al.*** (***2019***). HPV genotyping was performed for the samples at 1: before TCA treatment, 2: after TCA treatment, and 3: follow-up after TCA treatment.

### DNA extraction and 16S ribosomal RNA gene sequencing

LBC samples were sent to Tokai University (Isehara, Japan) for DNA extraction, amplification, and the 16S ribosomal RNA (rRNA) gene sequencing on an Ion PGM™ System (Thermo Fisher Scientific, Waltham, MA USA). The Ion 16S™ Metagenomics Kit was used. Single-end reads were generated for the seven hypervariable regions (V2, V3, V4, V6, V7, V8, and V9) ***Barb et al.*** (***2016***).

### Microbiome analysis

Microbiome bioinformatics was performed with R (version 4.2.1) and QIIME2-2020.11 ***Bolyen et al.*** (***2019***). Hereafter, the abbreviation q2-means QIIME 2 plugins and r-R packages. The Taxonomy database was built using q2-RESCRIPT and SILVA rRNA gene database (SILVA138.1) ***Robeson et al.*** (***2021***) ***Quast et al.*** (***2012***). We removed low-quality sequences, such as those containing five or more ambiguous bases and eight or more homopolymers. We adopted bacteria rRNA gene sequences over 1400 bp to build the taxonomy database ***Robeson et al.*** (***2021***) ***Yang et al.*** (***2016***). Naive-Bayes-based taxonomy classifier was created through q2-feature-classifier ***Bokulich et al.*** (***2018***) ***Pedregosa et al.*** (***2011***) ***Ziemski et al.*** (***2021***).

Demultiplexed 20 fastq files for samples before and after chemical peeling by TCA of 10 patients consisting of single-end reads were imported by q2-tools import command and visualized by q2-demux summarize. We generated an amplicon sequence variant (ASV) using q2-dada2 ***Callahan et al.*** (***2016***). The q2-dada2 denoised and dereplicated reads and removed chimeras ***Callahan et al.*** (***2016***). Using the option of q2-dada2, the reads were trimmed at 15 bp under no truncated and pooled options ***Pos*** (***2023***). The ASVs were assigned taxonomy through q2-feature-classifier classify-sklearn using above the Naive-Bayes-based taxonomy classifier ***Bokulich et al.*** (***2018***). The confidence parameter in q2-feature-classifier classify-sklearn was set as 0.5 to obtain a high-recall ***Bokulich et al.*** (***2018***). ASVs classified as Chloroplast, Mitochondria, Eukaryota, Unassigned and those with no phylum-level taxonomic information were removed by q2-taxa filter-table. As final quality filtering, we retained ASVs that had at least 90% identity and 90% query alignment with the SILVA ribosomal RNA gene database using q2-quality-control. The top 10 abundant taxa at the genus level were displayed in colored bar plots separated as before and after TCA treatment for each positive or negative value of high-risk HPV after TCA treatment ***McMurdie and Holmes*** (***2013***).

Species richness called alpha diversity for the difference within the sample was derived using q2-breakaway***Willis and Bunge*** (***2015***). The changes in alpha diversity before and after TCA treatment were plotted for each patient, divided into high-risk HPV positive and negative after TCA treatment. A test of similarity called beta diversity of the cervicovaginal microbiome composition pre- or post-TCA treatment was evaluated using the q2-diversity adonis***Anderson*** (***2001***) and plotted using the principal component analysis.

As differential abundance analysis for cervicovaginal microbiome before and after TCA treatment, we compared the abundance of taxa using DESeq2 in r-microbiomeMarker under the following parameter setting; normalization method: relative log expression; identity method: Wald; p-value adjustment for multiple comparisons: Benjamini-Hochberg; and p-value cutoff: 0.05***Cao et al.*** (***2022***)***Love et al.*** (***2014***).

## Data Availability

All data produced in the present study are available upon reasonable request to the authors.

## Acknowledgments

We would like to thank Editage (www.editage.com) for English language editing. We would like to thank the Medical Science College Office, Tokai University for technical assistance.

## Author contributions

To. S. is responsible for the study’s conception and design. Ta. S. and To. S. wrote and all authors revised the manuscript. Ta. S. conducted the analyses. O. A. and T. I. conducted DNA extraction and high-throughput sequencing of DNA. All authors interpreted the data.

